# Precision Colorectal Cancer Screening with Polygenic Risk Score

**DOI:** 10.1101/2020.08.19.20177931

**Authors:** Tõnis Tasa, Mikk Puustusmaa, Neeme Tõnisson, Berit Kolk, Peeter Padrik

## Abstract

Colorectal cancer (CRC) is the second most common cancer in women and third most common cancer in men. Genome-wide association studies have identified numerous genetic variants (SNPs) independently associated with CRC. The effects of such SNPs can be combined into a single polygenic risk score (PRS). Stratification of individuals according to PRS could be introduced to primary and secondary prevention. Our aim was to combine risk stratification of a sex-specific PRS model with recommendations for individualized CRC screening.

Previously published PRS models for predicting the risk of CRC were collected from the literature. These were validated on the UK Biobank (UKBB) consisting of a total of 458 696 quality-controlled genotypes with 1810 and 1348 prevalent male cases, and 2410 and 1810 incident male and female cases. The best performing sex-specific model was selected based on the AUC in prevalent data and independently validated in the incident dataset. Using Estonian CRC background information, we performed absolute risk simulations and examined the ability of PRS in risk stratifying individual screening recommendations. The best-performing model included 91 SNPs. The C-index of the best performing model in the dataset was 0.613 (SE = 0.007) and hazard ratio (HR) per unit of PRS was 1.53 (1.47 – 1.59) for males. Respective metrics for females were 0.617 (SE = 0.006) and 1.50 (1.44 – 1.58).

PRS risk simulations showed that a genetically average 50-year-old female doubles her risk by age 58 (55 in males) and triples it by age 63 (59 in males). In addition, the best performing PRS model was able to identify individuals in one of seven groups proposed by Naber et al. for different coloscopy screening recommendation regimens.

We have combined PRS-based recommendations for individual screening attendance. Our approach is easily adaptable to other nationalities by using population-specific background data of other genetically similar populations.

## Introduction

Colorectal cancer (CRC) is the second most common cancer in women and third most common cancer in men with an estimated 500 000 new cancer cases (1). Recent work has elucidated the importance of genetic events leading to colorectal cancer based on clinical and molecular studies of colorectal tumors (2, 3).

CRC prevalence is likely to grow in the future so it is necessary to develop approaches to prediagnosis screening (4). The current programs that use both invasive and non-invasive methods mostly target the population in ages 50–74 years (5, 6). General guidelines continue to evolve and current population screening in Europe is mostly performed using a fecal occult blood test (FOBT) and coloscopy (7) but participation remains quite low even in groups with a family history of CRC (8). In most countries that use the FOBT, screening is available in every 2 years. The screening schedule is less frequent with coloscopy and flexible sigmoidoscopy, generally every 10 years (9).

Colorectal cancer is known to have a significant genomic component with an estimated heritability of around 12–35% (10). Family-based studies have identified rare high-penetrance mutations in at least a dozen genes, but collectively, these account for only a small fraction of familial risk – 5% of colorectal cancers arise in the setting of a well-established Mendelian inherited disorder (11). Frequently described CRC-predisposition genes include *APC, MLH1, MSH2, MSH6, PMS2, STK11, MUTYH, SMAD4, BMPR1A, PTEN, TP53, CHEK2, POLD1, POLE* (12–14). Individuals with specific genetics linked hereditary CRC syndromes typically involve much more intense screening regimens often with annual coloscopy starting in young adulthood (15). Testing for genes associated with highly penetrant hereditary CRC syndromes is likely to provide significant clinical benefits in a cost-effective manner (16).

CRC risk factors have shown promise to stratify uniform screening programs (17, 18). Risk prediction models are attractive as they are non-invasive and are easier to implement in a general population or primary care screening setting. Known conventional risk factors include age, obesity, a diet high in fat and low in fibre, alcohol consumption, smoking, type II diabetes, and a family history of CRC (19). Published models include data routinely available from electronic health records such as age, gender, and body mass index (BMI), to more complex models containing detailed information about lifestyle factors and genetic biomarkers (20). QCancer10 (21) performs well in men, others include Tao *et al*. (2014) (22), Driver *et al*. (23), Ma *et al*. (2010) (24), Wells *et al*. (2014) (25). However, sporadic cancers derived from a larger number of common, low-penetrance genetic variants with individually small effects account for 70% of all CRC cases (26). Therefore, it would be important for risk stratification to apply information from the low penetrance genetic variants.

Genome-wide association studies (GWASs) for sporadic CRC have been extensively applied to identify independent signals associated with modifications in CRC risk (27–39). A recent study by Huyghe *et al*. enhanced the number of known independent signals for CRC to around 100 (11). Individual SNPs can be aggregated into a polygenic risk score (PRS). The combination of a large number of such SNPs in genetic risk scores has been demonstrated to enable relevant risk stratification (40–44). As such, PRS complements other factors that identify groups with modified risk and could serve as a stand-alone method for risk stratification before the diagnostic screening (45).

The aim of this study is to evaluate the CRC risk prediction performance of several published PRS and to assess its use as a risk stratification approach in the context of Estonia. Concretely, we aim to develop a model to input information into a PRS risk-stratified CRC screening regimen.

## Methods

### Participant data of UK Biobank

This study used genotypes from the UK Biobank cohort (obtained 07.11.2019) and made available to Antegenes under application reference number 53602. The data was collected, genotyped using either the UK BiLEVE or Affymetrix UK Biobank Axiom Array. Colorectal cancer cases in the UK Biobank cohort were retrieved by the status of ICD-10 codes C18, C19, and C20. We additionally included cases with self-reported UK Biobank code “1020”.

Quality control steps and in detail methods applied in imputation data preparation have been described by the UKBB and made available at http://www.ukbiobank.ac.uk/wp-content/uploads/2014/04/UKBiobank_genotyping_QC_documentation-web.pdf. We applied additional quality controls on autosomal chromosomes. First, we removed all variants with allele frequencies outside 0.1% and 99.9%, genotyping call rate < 0.1, imputation (INFO) score < 0.4 and Hardy-Weinberg equilibrium test p-value < 1E-6. Sample quality control filters were based on several pre-defined UK Biobank filters. We removed samples with excessive heterozygosity, individuals with sex chromosome aneuploidy, and excess relatives (> 10). Additionally, we only kept individuals for whom the submitted gender matched the inferred gender, and the genotyping missingness rate was below 5%.

Quality controlled samples were divided into prevalent and incident datasets for females and males separately. The prevalent datasets included CRC cases diagnosed before Biobank recruitment with 5 times as many controls without the diagnosis. Incident datasets included cases diagnosed in any of the linked databases after recruitment to the Biobank and all controls not included in the prevalent dataset.

Prevalent datasets were used for identifying the best sex-specific candidate model and the incident datasets were used to obtain an independent PRS effect estimate on CRC status.

### Model selection from candidate risk models

We searched the literature for PRS models in the public domain. The requirements for inclusion to the candidate set were the availability of the chromosomal location, reference and alternative allele, minor allele frequency, and an estimator for the effect size either as odds ratio (OR) or its logarithm (log-OR) specified for each genetic variant. In cases of iterative model developments on the same underlying base data, we retained chronologically newer ones. The search was performed with Google Scholar and PubMed web search engines by working through a list of publications using the search [“Polygenic risk score” or “genetic risk score” and “colorectal cancer”], and then manually checking the results for the inclusion criteria. We additionally pruned the PRS from multi-allelic, non-autosomal, non-retrievable variants based on bioinformatics re-analysis with Illumina GSA-24v1 genotyping.

PRSs were calculated as 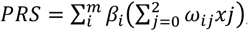 where *ω_ij_* is the probability of observing genotype *j*, where *j*∈*{0,1,2)* for the i-th SNP; *m* is the number of SNPs; and *β_i_* is the effect size of the i-th SNP estimated in the PRS. The mean and standard deviation of PRS in the cohort were extracted to standardize individual risk scores to Gaussian. We tested the assumption of normality with the mean of 1000 Shapiro-Wilks test replications on a random subsample of 1000 standardized PRS values.

Next, we evaluated the relationship between CRC status and standardized PRS in the two sex-specific prevalent datasets with a logistic regression model to estimate the logistic regression-based odds ratio per 1 standard deviation of PRS *(ORsd)*, its p-value, model Akaike information criterion (AIC) and Area Under the ROC Curve (AUC). The logistic regression model was compared to the null model using the likelihood ratio test and to estimate the Nagelkerke and McFadden pseudo-R^2^. We selected the candidate model with the highest AUC to independently assess risk stratification in the incident dataset.

### Independent performance evaluation of a polygenic risk score model

Firstly, we repeated the main analyses in the prevalent dataset. The main aim was to derive a primary risk stratification estimate, hazard ratio per 1 unit of standardized PRS (HR*sd)*, using a right-censored and left-truncated Cox-regression survival model. The start of time interval was defined as the age of recruitment; follow-up time was fixed as the time of diagnosis. Scaled PRS was fixed as the only independent variable of CRC diagnosis status. 95% confidence intervals were created using the standard error of the log-hazard ratio.

Further, we evaluated the concordance between theoretical hazard ratio estimates derived with the continuous per unit PRS (HRsd) estimate and the hazard ratio estimates inferred empirically from data. For this, we binned the individuals by PRS to 5%-percentiles and estimated the empiric hazard ratio of CRC directly between those classified in each bin and those within 40–60 PRS percentile. Theoretical estimates were derived from the relationship 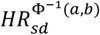, where the exponent is the expected Gaussian value between two arbitrary percentiles *a* and *b* (bounded between 0 and 1, a<b) of the Gaussian distribution, 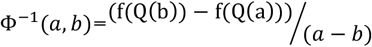, where *Q(b)* is the Gaussian quantile function on a percentile *b* and *f(Q(b))* is the Gaussian probability density function value at a quantile function value. We compared the two approaches by using the Spearman correlation coefficient and the proportion of distribution-based 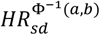 estimates in empirical confidence intervals.

### Absolute risk estimation

Individual *τ*-year (eg. 10-year) absolute risk calculations are based on the risk model developed by Pal Choudhury *et al* (46). Individual absolute risks are estimated for currently *a*-year old individuals in the presence of known risk factors *(Z)* and their relative log hazard-ratio parameters (*β*). 95% uncertainty intervals for the hazard ratio were derived using the standard error and z-statistic 95% quantiles CI_HR=exp_(*β*±1.96*se(HRsd)), where se(HRsd) is the standard error of the log-hazard ratio estimate. Risk factors have a multiplicative effect on the baseline hazard function. The model specifies the next *τ*-year absolute risk for a currently *a*-year old individual as

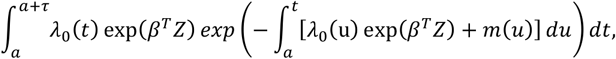

where *m(t)* is age-specific mortality rate function and *λ*_0_*(t) is* the baseline-hazard function, *t* ≥ *T* and *T* is the time to onset of the disease. The baseline-hazard function is derived from marginal age-specific CRC incidence rates (*λ_m_(t))* and distribution of risk factors *Z* in the general population *(F(z))*.

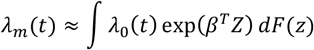

This absolute risk model allows disease background data from any country. In this analysis, we used Estonian background information. We calculated average cumulative risks using data from the National Institute of Health Development of Estonia (47) that provides population average disease rates in age groups of 5-year intervals. Sample sizes for each age group were acquired from Statistics Estonia for 2013–2016. Next, we assumed constant incidence rates for each year in the 5-year groups. Thus, incidence rates for each age group were calculated as *IR* = *X_t_/N_t_*, where X_t_ is the number of first-time cases at age *t* and N_*t*_ is the total number of individuals in this age group. Final per-year incidences were averaged over time range 2013–2016. Age-and sex-specific mortality data were retrieved from the World Health Organization (48) and competing mortality rates were constructed by subtracting yearly age- and sex-specific disease mortality rates from general mortality rates. Colorectal cancer mortality estimates were derived from the Global Cancer Observatory (49).

We applied this model to estimate absolute risks for individuals in the 1^st^, 10^th^, 25^th^, 50^th^, 75^th^, 90^th^ and 99^th^ PRS quantiles, eg. an individual on the 50^th^ percentile would have a standardized PRS of 0. Confidence intervals for the absolute risk are estimated with the upper and lower confidence intervals of the continuous per unit log-hazard ratio. Similarly, we used the absolute risk model to estimate lifetime risks (between ages 0 and 85) for the individuals in the same risk percentiles.

### A polygenic risk score based screening recommendations

Next, we simulate cumulative PRS stratified lifetime risks using sex-specific CRC incidences in the Estonian population by evaluating lifetime absolute risks for individuals in various PRS risk percentiles. Additionally, we assessed the differences in ages where individuals in various PRS risk percentiles attain 1 to 3-fold increases of risk compared to the 10-year risk of an average individual of the same sex.

Lastly, we combine PRS risk-based screening recommendations from Naber *et al*. (50) with PRS-based relative risk. They optimized the screening intervals against a hypothetical cost scenario under various AUC levels of PRS models. We adapt these recommendations to support relative risks and estimate the proportion of individuals given different screening recommendations with our best performing PRS model for a basis to provide individualized CRC screening recommendations. We use the ratio between the population stratified with our best performing PRS and relative risks of 10-year CRC to estimate the proportion of individuals in each available recommendation category.

## Results

### Polygenic risk score re-validation in a UKBB population cohort

There was a total of 487,410 quality-controlled genotypes available for males and females in the complete UK Biobank cohort. Phenotype data was available for a total of 458 696 individuals. This included 242 832 (241 022 controls, 1810 cases) cases for incident females and 6230 (5198 controls, 1032 cases) for prevalent females, and additional 8117 prevalent (6769 controls, 1348 cases) males and 201 517 (199 107 controls, 2410 cases) incident males.

Altogether, 5 PRS models from 3 different publications were revalidated (11, 40, 51). Three different models adapted from Huyghe *et al*. included different subsets of identified variants: CRC7 was composed of loci previously reported at genome-wide significance, CRC4 of the variants presented in the meta-analysis of known and novel CRC risk loci and CRC6 was combined of variants that the authors used in PRS analyses. Normality assumption of the standardized PRS was not violated with any tested models (Shapiro-Wilks test p-values in CRC3 = 0.36, CRC4 = 0.48, CRC5 = 0.48, CRC6 = 0.49, CRC7 = 0.48). The best performing model was selected based on AUC, ORsd,, AIC and pseudo-R^2^ metrics for both females and males. The CRC6 model that was based on Huyghe et al. (11) performed the best (Table 1). The model’s AUC under the ROC curve for the association between the PRS and CRC diagnosis was 0.626 (SE = 0.018) for males (Figure 1) and 0.622 (SE = 0.021) for females.

**Table 1.**
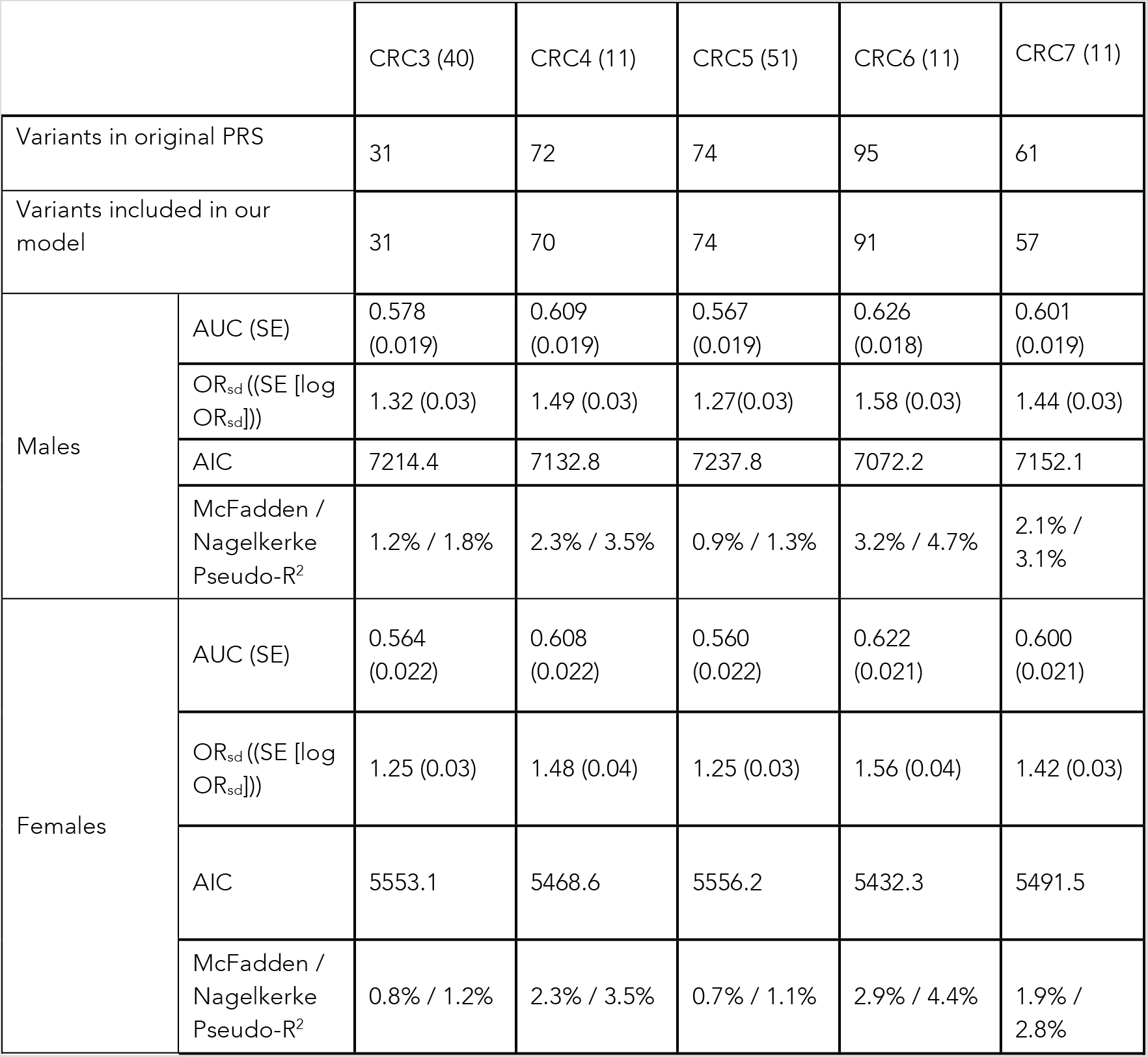
Comparison metrics of colorectal cancer PRS models based on the prevalent *UK Biobank* dataset.

|  |  | CRC3 (40) | CRC4 (11) | CRC5 (51) | CRC6 (11) | CRC7 (11) |
| --- | --- | --- | --- | --- | --- | --- |
| Variants in original PRS |  | 31 | 72 | 74 | 95 | 61 |
| Variants included in our model |  | 31 | 70 | 74 | 91 | 57 |
| Males | AUC (SE) | 0.578<br>(0.019) | 0.609<br>(0.019) | 0.567<br>(0.019) | 0.626<br>(0.018) | 0.601<br>(0.019) |
| | $OR_{sd}$ ((SE [log $OR_{sd}$])) | 1.32 (0.03) | 1.49 (0.03) | 1.27(0.03) | 1.58 (0.03) | 1.44 (0.03) |
|  | AIC | 7214.4 | 7132.8 | 7237.8 | 7072.2 | 7152.1 |
| | McFadden / Nagelkerke Pseudo-$R^2$ | 1.2% / 1.8% | 2.3% / 3.5% | 0.9% / 1.3% | 3.2% / 4.7% | 2.1% / 3.1% |
| Females | AUC (SE) | 0.564<br>(0.022) | 0.608<br>(0.022) | 0.560<br>(0.022) | 0.622<br>(0.021) | 0.600<br>(0.021) |
| | $OR_{sd}$ ((SE [log $OR_{sd}$])) | 1.25 (0.03) | 1.48 (0.04) | 1.25 (0.03) | 1.56 (0.04) | 1.42 (0.03) |
|  | AIC | 5553.1 | 5468.6 | 5556.2 | 5432.3 | 5491.5 |
| | McFadden / Nagelkerke Pseudo-$R^2$ | 0.8% / 1.2% | 2.3% / 3.5% | 0.7% / 1.1% | 2.9% / 4.4% | 1.9% / 2.8% |

**Figure 1.**
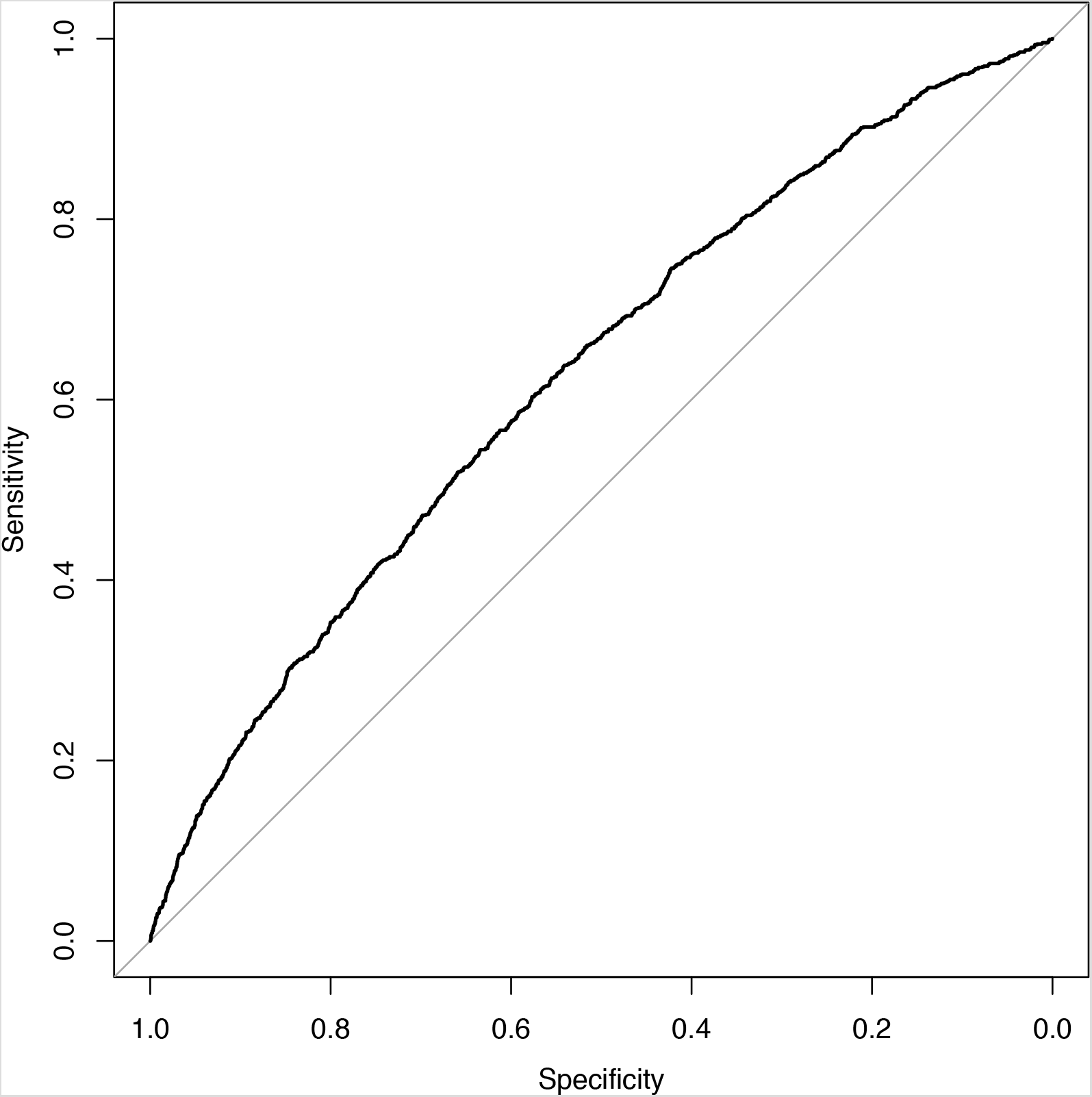
ROC plot of CRC cases and controls in the UK Biobank prevalent dataset of males.

Next, we evaluated the performance of the best performing CRC6 model in an independent UK Biobank incident dataset with the main aim of estimating the hazard ratio per unit of PRS. Table 2 presents the performance estimation metrics in the incident dataset. Hazard ratio per 1 unit of standard deviation (HR_sd_) in model CRC6 was *1.53* with standard error (log (*HR*)) = 0.02) for males. The concordance index (C-index) of the survival model testing the relationship between PRS and CRC diagnosis status in the females’ dataset was 0.617 (0.006) and highly similar in males.

**Table 2.**
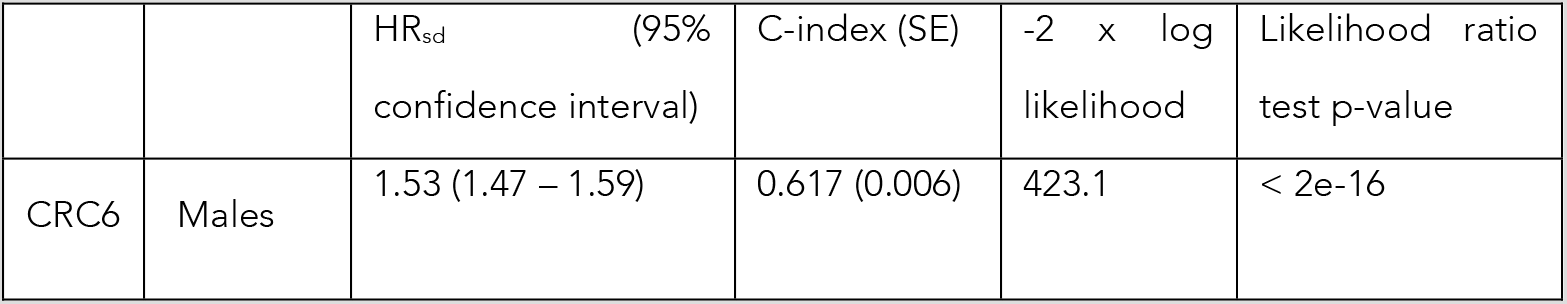
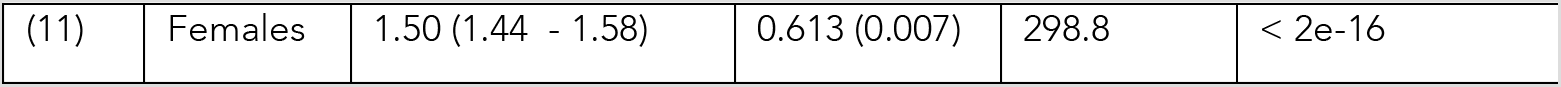
Performance metrics of the CRC6 model in the incident UK Biobank dataset.

| | | $HR_{sd}$ (95% confidence interval) | C-index (SE) | $-2 \times \log$ likelihood | Likelihood ratio test p-value |
| --- | --- | --- | --- | --- | --- |
| CRC6 | Males | 1.53 (1.47 – 1.59) | 0.617 (0.006) | 423.1 | < 2e-16 |
| (11) | Females | 1.50 (1.44 - 1.58) | 0.613 (0.007) | 298.8 | < 2e-16 |

Hazard ratio estimates compared to individuals in 40–60 percentile of PRS for females and males are visualized on Figure 2. For both females (panel A) and males (panel B), the theoretical hazard ratio matched in empirical estimate’s confidence intervals in 16 out of 16 comparisons. Alternatively, the Spearman correlation coefficient between the empiric and theoretical hazard ratio estimates was 0.992 for males and 0.995 for females indicating near-perfect correspondence.

**Figure 2.**
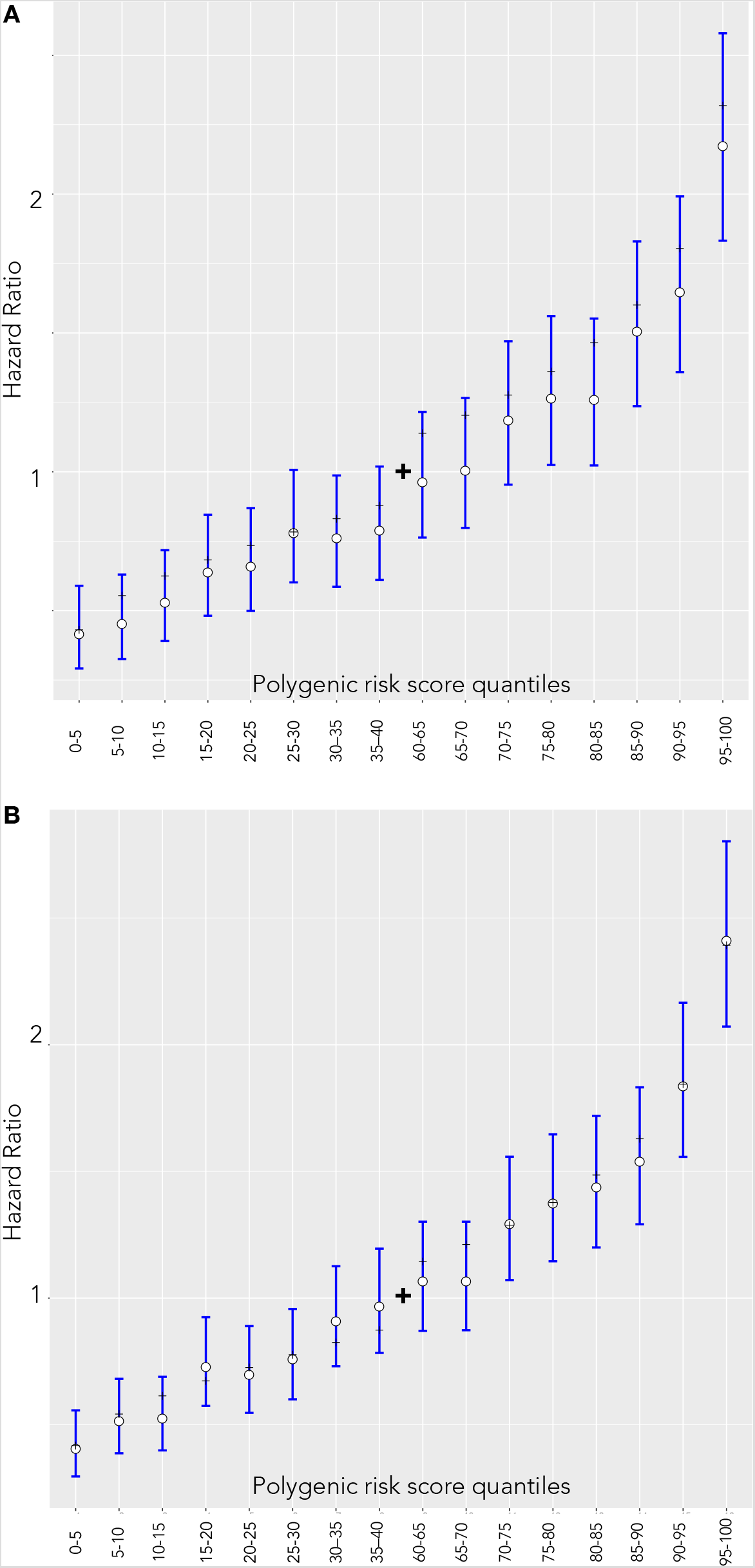
Hazard ratio estimates between quantiles 40–60 of the PRS and categorized 5% bins in the UKBB incident dataset. White dots and blue lines represent empirically estimated hazard ratio estimates and corresponding confidence intervals. Black dashes represent the theoretical hazard ratio for the 5%-quantile bins derived from the hazard ratio of per unit PRS. (A) Females (B) Males

### Polygenic risk score in colorectal cancer screening stratification

We used a model by Pal Choudhury et al. to derive individual 10-year risks (46) and specified *F(z)* as the distribution of PRS estimates in the whole UKBB cohort. The log-hazard ratio (*β*) is based on the sex-specific estimate of the log-hazard ratio in the CRC6 model of the incident UKBB dataset. Age-specific CRC incidence and competing mortality rates provided the background for CRC incidences in the Estonian population.

In the Estonian population, the absolute risk of developing colorectal cancer in the next 10 years among 50-year old men in the 1^st^ percentile is 0.16% (0.14% – 0.18%) and 1.15% (1.07% – 1.24%) in the 99^th^ percentile (0.16% [0.14% – 0.18%] and 1.06% [0.97% – 1.16%] for women, respectively). At age 70, corresponding risks for the same percentiles of men become 1.07% (0.96% – 1.19%) and 7.4% (6.85% –7.96%). Considerably lower estimates were found for women: 0.71% (0.63% – 0.81%) and 4.67% (4.27% –5.09%) respectively. The relative risks between the most extreme percentiles are therefore around 6.7-fold. Similarly, competing risks accounted cumulative risks of females in the 99th percentile surpass 11.4% (10.6% – 12.2%) by age 85 but remain at 1.71% (1.53% – 1.91%) in the 1^st^ percentile (Figure 3). Equivalent values for women are somewhat lower: 10.2% (9.33% – 11.1%) and 1.6% (1.41% – 2.6%).

**Figure 3.**
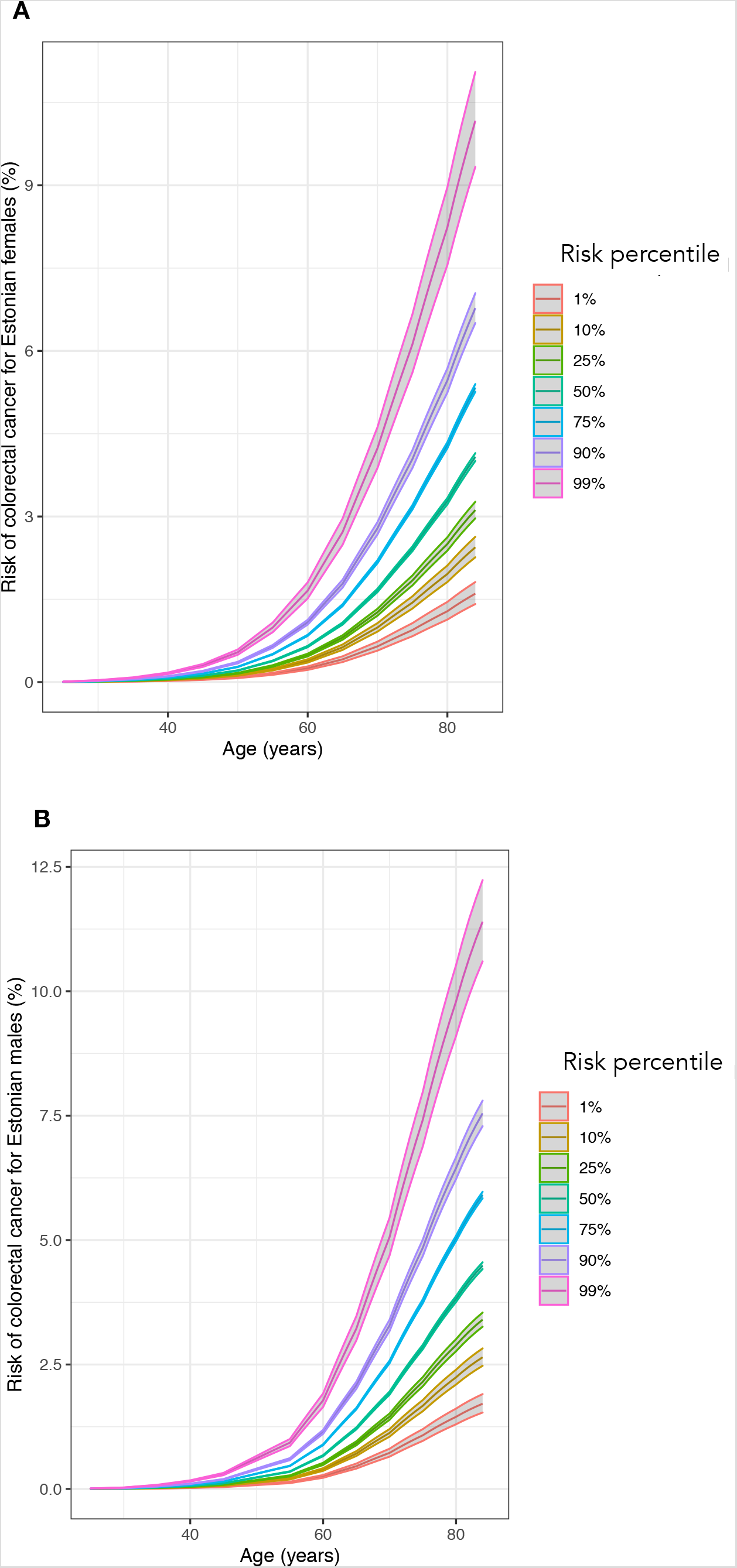
Cumulative risks (%) of colorectal cancer between ages 20 to 85 in various risk percentiles. (A) Females (B) Males

Genetically average 50-year-old males have a 10-year risk of CRC equaling 0.432% and for females it is 0.412%. A 41-year-old male in the 99^th^ percentile (42 in females) has a larger risk than an average 50-year-old. At the same time, males in the 1^st^ percentile attain this risk by age 58 (61 in females). Males above the 95^th^ percentile (96^th^ percentile in females) have a more than 2-fold risk increase compared to the average. A genetically average female doubles her risk at age 50 by age 58 (55 in males) and triples it by age 63 (59 in males). The 1^st^ percentile of females only attains the double of average 50-year old’s risk by age 72 (66 in males) (Figure 4).

**Figure 4.**
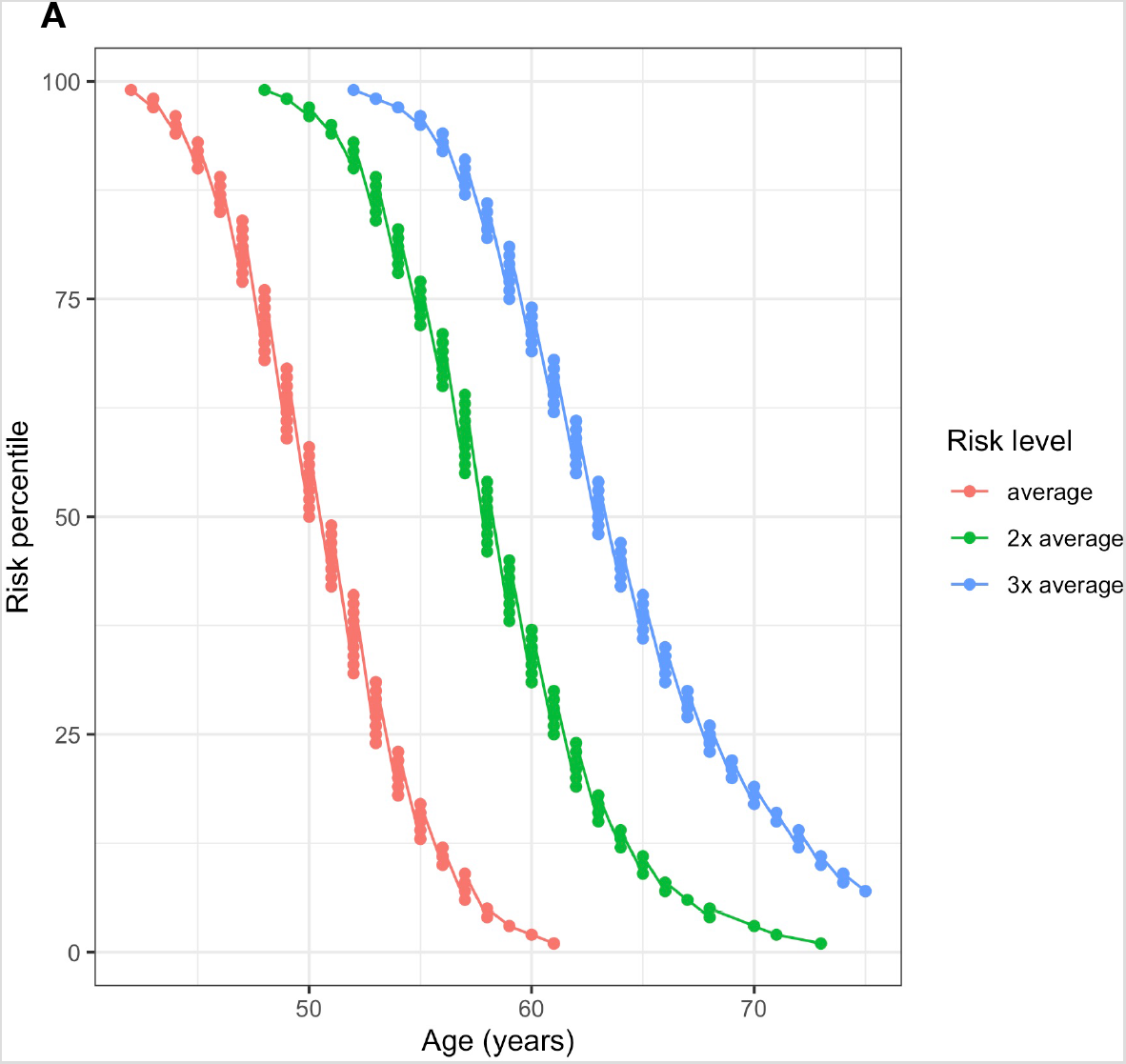
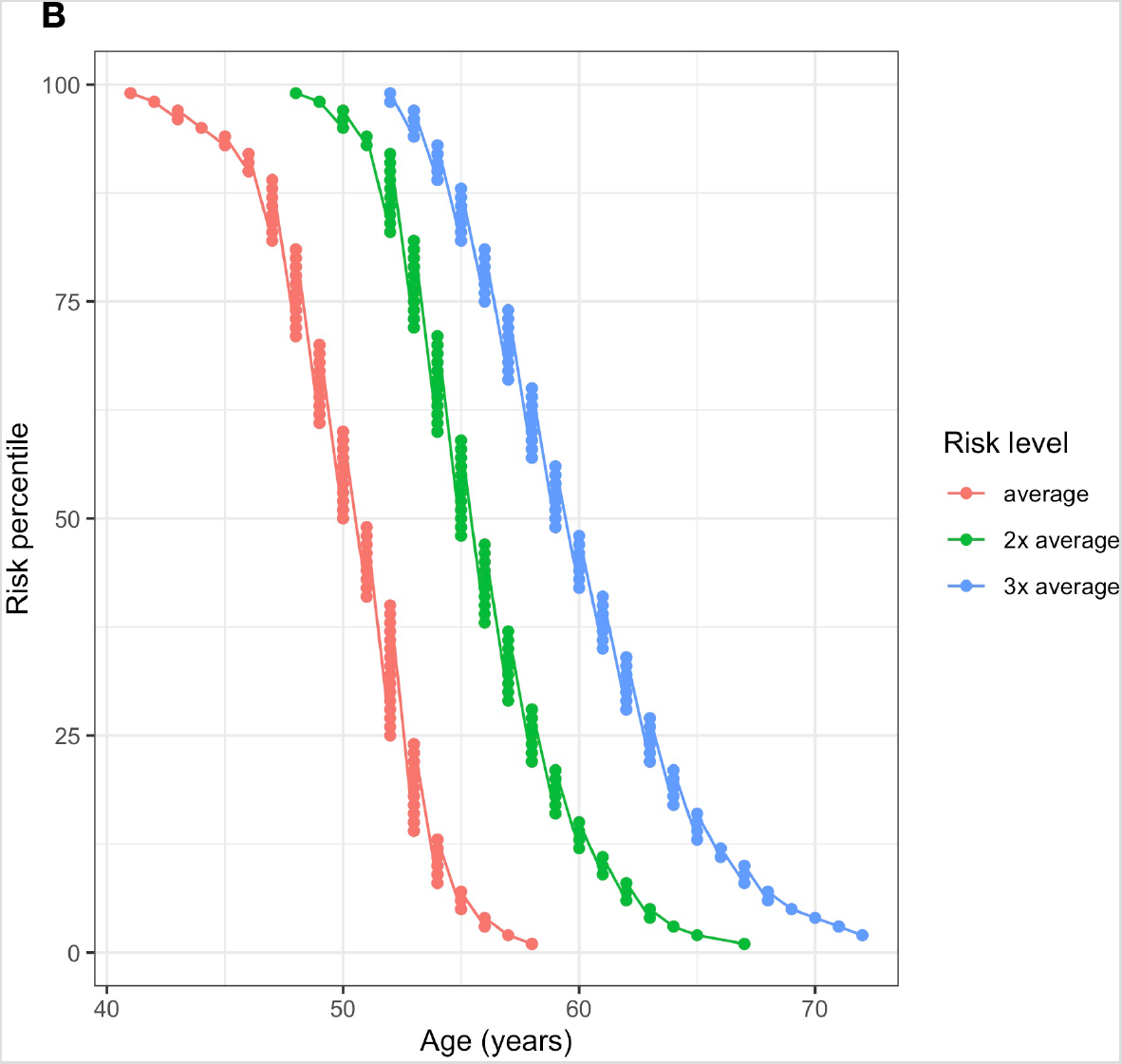
Ages when Estonian individuals in different risk percentiles attain 1–3 fold multiples of 10-year risk compared to 50-year old individuals with population average PRS (Risk level: “average”). (A) Females (B) Males

Relative risk stratification with PRS provided a basis for personal CRC screening intervals. Recommendations for coloscopy based screening presented below are adapted from Naber et al. (50) for a model with AUC = 0.6 and required individual relative risks as input. CRC screening is individualized by differences in the intervals of planned coloscopies. Relative risks are derived from 10-year risk differences compared to average PRS. As an alternative to coloscopy, we recommend annual fecal immunochemical testing, with individualized starting from the age at which the patient attains the 10-year risk of the average 50-year-old person (52–54).

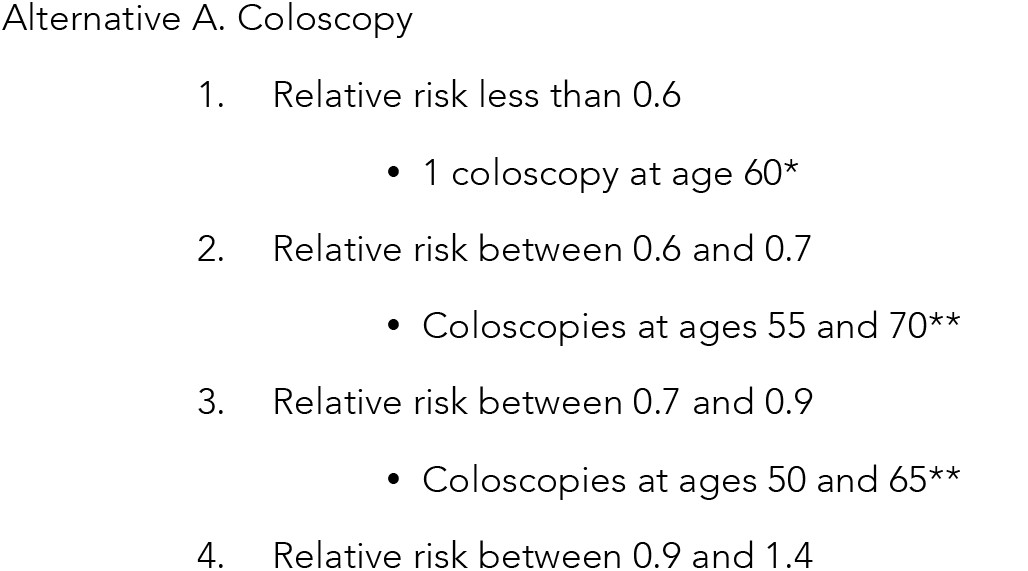
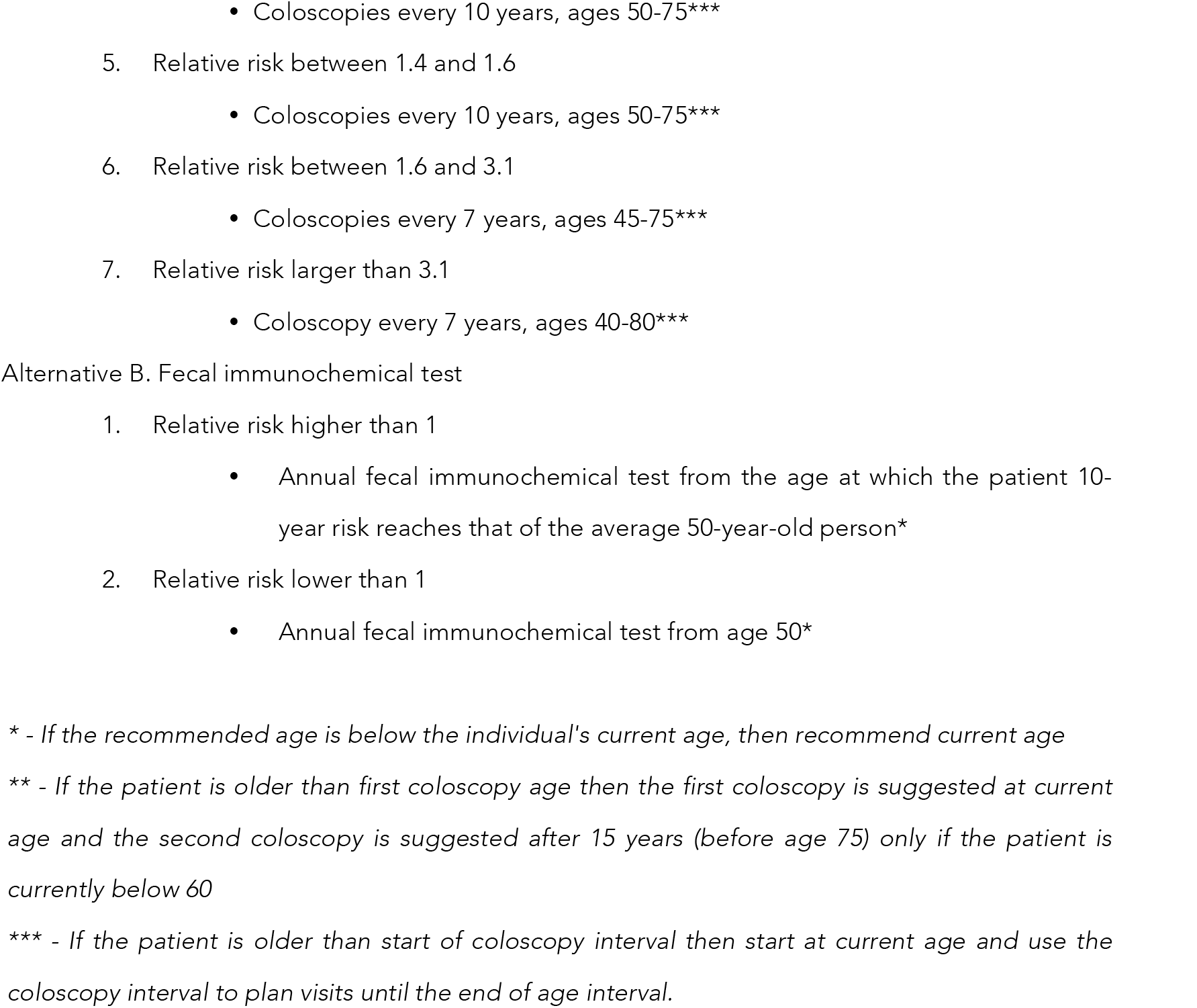

Next, we estimated the proportion of individuals with relative risks to an average individual in the Estonian population. We estimated that relative risks as more than 3.1, compared to an individual with median population PRS, in around 0.3% of females, between 1.6 and 3.1 in 11.9%, between 1.4 and 1.6 in 10.3%, between 0.9 and 1.4 in 37.7%, between 0.7 and 0.9 in 20.8%, between 0.6 and 0.7 in 9.4% and below 0.6 in 9.5%. The equivalent proportions for males were highly similar.

## Discussion

In this study, we validated different publicly available PRS models to find a model to use for individualizing screening recommendations. Several studies have previously explored various aspects of PRS performances. Huyghe *et al*. (11) performed a combined meta-analysis to identify around 40 new independent signals at *P*< 5×10^−8^ and derived a PRS derived with 95 independent association signals. They also demonstrated the application of PRS to preventive screening by simulating the start of screening ages based on 10-year average risks. Iwasaki *et al*. constructed a PRS with AUC = 0.63 that was better than a non-genetic risk prediction score (AUC = 0.60) that included age, body mass index, and tobacco and alcohol use. AUC of a combined model improved to 0.66. (55). Weigl *et al*. demonstrated that the PRS effectively discriminated between the risk for advanced neoplasms versus normal coloscopy findings (56). The study estimated that participants with the highest tertile of PRS have the same risk of advanced colorectal neoplasm as participants 17.5 years older from the lowest tertile of PRS. In a different study by Weigl *et al*., a 53 SNP PRS and family history of CRC were both associated with increased CRC risk but appeared to be independent of one another (57). PRS may augment family history–based CRC risk stratification. Archambault *et al*. have demonstrated the utility of using PRS for detecting the risk of early-onset CRC as opposed to late-onset. Comparing the higher versus the lowest PRS quartile, the risk increased 3.7-fold and 2.9-fold for early- and late-onset CRC, respectively. Participants without a first-degree family history of CRC had the strongest association. Comparing the highest with the lowest quartiles in this group, the risk increased 4.3-fold versus 2.9-fold for early- versus late-onset CRC (58). Along with lifestyle and environmental risk profiling, individuals at increased susceptibility to early-onset CRC may be used to design personalized screening regimens for high-risk individuals < 50 years of age (58). Schmit et al. identified around 4.3% of the population with a greater odds ratio than 2 for developing CRC (51). Again, these results corroborate the overall risk stratification observed in current model re-validation.

Screening of CRC aims to lead to early diagnosis and more effective treatment by detecting and removing premalignant polyps before they progress to CRC (59). It substantially reduces the incidence and mortality of the disease (60). Across tools available for screening the numbers of CRC deaths prevented appear to be relatively similar, although sensitivity and specificity for detection of polyps and adenomas of CRC vary. Studies indicate that coloscopy, sigmoidoscopy, and both guaiac-based FOBT and fecal immunochemical tests are associated with a decreased risk of CRC mortality. The effectiveness of other methods such as multi-targeted stool DNA testing is better than FOBT due to greater sensitivity for advanced adenomas and early CRC (61). The invasive methods, such as coloscopy, have the highest sensitivity for CRC and adenomas and enable the detection and removal of precancerous lesions (62). Coloscopy is the preferred test for patients at higher risk for CRC due to having either one first degree relative with CRC or documented advanced adenoma or serrated lesion at age < 60 years, or two or more first-degree relatives with such findings at any age (63). However, the uncomfortable and invasive nature of the procedure contributes to poor patient compliance.

Risk factor-based models have been used to test different screening set-up modalities. Stanesby *et al*. compared the expected colorectal cancer deaths under three screening programs; age-based, genetic-based and combined age-based and genetic-based. They found that screening eligibility based on the genetic risk profile for age is as efficient as eligibility based on age alone for preventing colorectal cancer mortality, but identifies an additional 7% of the population at sufficient risk to benefit from screening who would not normally be screened given they are aged under 50 years (3). Weigl et al. studied of PRS can identify individuals with clinically relevant neoplasms in a screening coloscopy population (56). They found that increased PRS was associated with a higher prevalence of advanced neoplasms as a successful use in a CRC screening study (56). This supports the use of genetic data in population screening stratification.

Naber et al. (50) optimized screening intervals for cost-efficiency and found that screening based on stratification with a baseline model with AUC of 0.6 would become cost-effective with PRS prices below 141 US dollars. Here, we have adapted their screening recommendations to a PRS model from Huyghe et al. adapted and locally revalidated for individual (11). In addition, based on international guidelines, an alternative option to coloscopy is the annual fecal immunochemical test from the age at which the risk of the average 50-year-old person is reached. This combination allows developing inputs for individualized PRS based individual screening recommendations.

In conclusion, we have combined and adapted a PRS based model for use with PRS informed screening recommendations. Our PRS model performed equally well for males and females identifying individuals from seven different screening setups. The genetic risk-based recommendations can be applied prospectively by individuals and also by institutions aiming to make screening provision more efficient.

## Data Availability

Individual level genotype and phenotype data from UK Biobank can not be explicitly shared. The UK Biobank Resource was used under Application Reference Number 53602. New users can request access
to UK Biobank from http://www.ukbiobank.ac.uk/resources/.

## Acknowledgments

This research has been conducted using the UK Biobank Resource under Application Reference Number 53602 and with support by EIT Health the Digital Sandbox program.

## Ethical approval

### UK Biobank

The UK Biobank study was approved by the North West Multi-Centre Research Ethics Committee (UK Biobank reference: 16/NW/0274). All participants provided written informed consent to participate in the UK Biobank study.

## Statement of data availability

Individual level genotype and phenotype data from UK Biobank can not be explicitly shared. The UK Biobank Resource was used under Application Reference Number 53602. New users can request access to UK Biobank from http://www.ukbiobank.ac.uk/resources/.

## Funding

OÜ Antegenes has received a grant from the EIT Health The Digital Sandbox program and additional Innovation Voucher funding meant for business development of small and medium sized Estonian enterprises.

## Notes

### Competing Interest Statement

The authors have declared no competing interest.

### Funding Statement

OU Antegenes has received a grant from the EIT Health The Digital Sandbox program and additional Innovation Voucher funding meant for business development of small and medium sized Estonian enterprises.

### Author Declarations

UK Biobank: The UK Biobank study was approved by the North West Multi-Centre Research Ethics Committee (UK Biobank reference: 16/NW/0274). All participants provided written informed consent to participate in the UK Biobank study.

## References

1. Ferlay J, Colombet M, Soerjomataram I, Dyba T, Randi G, Bettio M, et al. Cancer incidence and mortality patterns in Europe: Estimates for 40 countries and 25 major cancers in 2018. European journal of cancer (Oxford, England: 1990). 2018;103:356–87.

2. Genetic testing for colon cancer: joint statement of the American College of Medical Genetics and American Society of Human Genetics. Joint Test and Technology Transfer Committee Working Group. Genetics in medicine: official journal of the American College of Medical Genetics. 2000;2(6):362–6.

3. Stanesby O, Jenkins M. Comparison of the efficiency of colorectal cancer screening programs based on age and genetic risk for reduction of colorectal cancer mortality. European journal of human genetics: EJHG. 2017;25(7):832–8.

4. Arnold M, Sierra MS, Laversanne M, Soerjomataram I, Jemal A, Bray F. Global patterns and trends in colorectal cancer incidence and mortality. Gut. 2017;66(4):683–91.

5. Senore C, Basu P, Anttila A, Ponti A, Tomatis M, Vale DB, et al. Performance of colorectal cancer screening in the European Union Member States: data from the second European screening report. Gut. 2018.

6. Schreuders EH, Ruco A, Rabeneck L, Schoen RE, Sung JJ, Young GP, et al. Colorectal cancer screening: a global overview of existing programmes. Gut. 2015;64(10):1637–49.

7. Zavoral M, Suchanek S, Zavada F, Dusek L, Muzik J, Seifert B, et al. Colorectal cancer screening in Europe. World journal of gastroenterology: WJG. 2009;15(47):5907.

8. Tsai M-H, Xirasagar S, Li Y-J, De Groen PC. Peer Reviewed: Colonoscopy Screening Among US Adults Aged 40 or Older With a Family History of Colorectal Cancer. Preventing chronic disease. 2015;12.

9. Rex DK. Screening Tests for Colon Cancer. Gastroenterology & hepatology. 2016;12(3):197–9.

10. Mucci LA, Hjelmborg JB, Harris JR, Czene K, Havelick DJ, Scheike T, et al. Familial Risk and Heritability of Cancer Among Twins in Nordic Countries. Jama. 2016;315(1):68–76.

11. Huyghe JR, Bien SA, Harrison TA, Kang HM, Chen S, Schmit SL, et al. Discovery of common and rare genetic risk variants for colorectal cancer. Nature genetics. 2019;51(1):76–87.

12. Fishel R, Lescoe MK, Rao MR, Copeland NG, Jenkins NA, Garber J, et al. The human mutator gene homolog MSH2 and its association with hereditary nonpolyposis colon cancer. Cell. 1993;75(5):1027–38.

13. Bronner CE, Baker SM, Morrison PT, Warren G, Smith LG, Lescoe MK, et al. Mutation in the DNA mismatch repair gene homologue hMLH1 is associated with hereditary non-polyposis colon cancer. Nature. 1994;368(6468):258–61.

14. Al-Tassan N, Chmiel NH, Maynard J, Fleming N, Livingston AL, Williams GT, et al. Inherited variants of MYH associated with somatic G:C-->T:A mutations in colorectal tumors. Nature genetics. 2002;30(2):227–32.

15. Syngal S, Brand RE, Church JM, Giardiello FM, Hampel HL, Burt RW. ACG clinical guideline: genetic testing and management of hereditary gastrointestinal cancer syndromes. The American journal of gastroenterology. 2015;110(2):223.

16. Gallego CJ, Shirts BH, Bennette CS, Guzauskas G, Amendola LM, Horike-Pyne M, et al. Next-Generation Sequencing Panels for the Diagnosis of Colorectal Cancer and Polyposis Syndromes: A Cost-Effectiveness Analysis. Journal of clinical oncology: official journal of the American Society of Clinical Oncology. 2015;33(18):2084–91.

17. Win AK, Macinnis RJ, Hopper JL, Jenkins MA. Risk prediction models for colorectal cancer: a review. Cancer epidemiology, biomarkers & prevention: a publication of the American Association for Cancer Research, cosponsored by the American Society of Preventive Oncology. 2012;21(3):398–410.

18. Read TE, Kodner IJ. Colorectal cancer: risk factors and recommendations for early detection. American Family Physician. 1999;59(11):3083.

19. Le Marchand L, Wilkens LR, Kolonel LN, Hankin JH, Lyu L-C. Associations of sedentary lifestyle, obesity, smoking, alcohol use, and diabetes with the risk of colorectal cancer. Cancer research. 1997;57(21):4787–94.

20. Usher-Smith JA, Harshfield A, Saunders CL, Sharp SJ, Emery J, Walter FM, et al. External validation of risk prediction models for incident colorectal cancer using UK Biobank. British journal of cancer. 2018;118(5):750–9.

21. Hippisley-Cox J, Coupland C. Development and validation of risk prediction algorithms to estimate future risk of common cancers in men and women: prospective cohort study. BMJ open. 2015;5(3):e007825.

22. Sharara AI, Harb AH. Development and validation of a scoring system to identify individuals at high risk for advanced colorectal neoplasms who should undergo colonoscopy screening. Clinical gastroenterology and hepatology: the official clinical practice journal of the American Gastroenterological Association. 2014;12(12):2135–6.

23. Driver JA, Gaziano JM, Gelber RP, Lee IM, Buring JE, Kurth T. Development of a risk score for colorectal cancer in men. The American journal of medicine. 2007;120(3):257–63.

24. Ma E, Sasazuki S, Iwasaki M, Sawada N, Inoue M. 10-Year risk of colorectal cancer: development and validation of a prediction model in middle-aged Japanese men. Cancer epidemiology. 2010;34(5):534–41.

25. Wells BJ, Kattan MW, Cooper GS, Jackson L, Koroukian S. Colorectal cancer predicted risk online (CRC-PRO) calculator using data from the multi-ethnic cohort study. Journal of the American Board of Family Medicine: JABFM. 2014;27(1):42–55.

26. Fearon ER, Vogelstein B. A genetic model for colorectal tumorigenesis. Cell. 1990;61(5):759–67.

27. Broderick P, Carvajal-Carmona L, Pittman AM, Webb E, Howarth K, Rowan A, et al. A genome-wide association study shows that common alleles of SMAD7 influence colorectal cancer risk. Nature genetics. 2007;39(11):1315–7.

28. Tomlinson I, Webb E, Carvajal-Carmona L, Broderick P, Kemp Z, Spain S, et al. A genome-wide association scan of tag SNPs identifies a susceptibility variant for colorectal cancer at 8q24.21. Nature genetics. 2007;39(8):984–8.

29. Zanke BW, Greenwood CM, Rangrej J, Kustra R, Tenesa A, Farrington SM, et al. Genome-wide association scan identifies a colorectal cancer susceptibility locus on chromosome 8q24. Nature genetics. 2007;39(8):989–94.

30. Berndt SI, Potter JD, Hazra A, Yeager M, Thomas G, Makar KW, et al. Pooled analysis of genetic variation at chromosome 8q24 and colorectal neoplasia risk. Human molecular genetics. 2008;17(17):2665–72.

31. Houlston RS, Webb E, Broderick P, Pittman AM, Di Bernardo MC, Lubbe S, et al. Meta-analysis of genome-wide association data identifies four new susceptibility loci for colorectal cancer. Nature genetics. 2008;40(12):1426–35.

32. Jaeger E, Webb E, Howarth K, Carvajal-Carmona L, Rowan A, Broderick P, et al. Common genetic variants at the CRAC1 (HMPS) locus on chromosome 15q13.3 influence colorectal cancer risk. Nature genetics. 2008;40(1):26–8.

33. Tenesa A, Farrington SM, Prendergast JG, Porteous ME, Walker M, Haq N, et al. Genome-wide association scan identifies a colorectal cancer susceptibility locus on 11q23 and replicates risk loci at 8q24 and 18q21. Nature genetics. 2008;40(5):631–7.

34. Tomlinson IP, Webb E, Carvajal-Carmona L, Broderick P, Howarth K, Pittman AM, et al. A genome-wide association study identifies colorectal cancer susceptibility loci on chromosomes 10p14 and 8q23.3. Nature genetics. 2008;40(5):623–30.

35. Houlston RS, Cheadle J, Dobbins SE, Tenesa A, Jones AM, Howarth K, et al. Meta-analysis of three genome-wide association studies identifies susceptibility loci for colorectal cancer at 1q41, 3q26.2, 12q13.13 and 20q13.33. Nature genetics. 2010;42(11):973–7.

36. Tomlinson IP, Carvajal-Carmona LG, Dobbins SE, Tenesa A, Jones AM, Howarth K, et al. Multiple common susceptibility variants near BMP pathway loci GREM1, BMP4, and BMP2 explain part of the missing heritability of colorectal cancer. PLoS genetics. 2011;7(6):e1002105.

37. Peters U, Jiao S, Schumacher FR, Hutter CM, Aragaki AK, Baron JA, et al. Identification of Genetic Susceptibility Loci for Colorectal Tumors in a Genome-Wide Meta-analysis. Gastroenterology. 2013;144(4):799–807.e24.

38. Whiffin N, Hosking FJ, Farrington SM, Palles C, Dobbins SE, Zgaga L, et al. Identification of susceptibility loci for colorectal cancer in a genome-wide meta-analysis. Human molecular genetics. 2014;23(17):4729–37.

39. Al-Tassan NA, Whiffin N, Hosking FJ, Palles C, Farrington SM, Dobbins SE, et al. A new GWAS and meta-analysis with 1000 Genomes imputation identifies novel risk variants for colorectal cancer. Scientific reports. 2015;5:10442.

40. Hsu L, Jeon J, Brenner H, Gruber SB, Schoen RE, Berndt SI, et al. A model to determine colorectal cancer risk using common genetic susceptibility loci. Gastroenterology. 2015;148(7):1330–9.e14.

41. Jenkins MA, Makalic E, Dowty JG, Schmidt DF, Dite GS, MacInnis RJ, et al. Quantifying the utility of single nucleotide polymorphisms to guide colorectal cancer screening. Future oncology (London, England). 2016;12(4):503–13.

42. Zhang B, Shrubsole MJ, Li G, Cai Q, Edwards T, Smalley WE, et al. Association of genetic variants for colorectal cancer differs by subtypes of polyps in the colorectum. Carcinogenesis. 2012;33(12):2417–23.

43. Abuli A, Castells A, Bujanda L, Lozano JJ, Bessa X, Hernandez C, et al. Genetic Variants Associated with Colorectal Adenoma Susceptibility. PloS one. 2016;11(4):e0153084.

44. Xin J, Chu H, Ben S, Ge Y, Shao W, Zhao Y, et al. Evaluating the effect of multiple genetic risk score models on colorectal cancer risk prediction. Gene. 2018;673:174–80.

45. Frampton MJ, Law P, Litchfield K, Morris EJ, Kerr D, Turnbull C, et al. Implications of polygenic risk for personalised colorectal cancer screening. Annals of oncology: official journal of the European Society for Medical Oncology. 2016;27(3):429–34.

46. Pal Choudhury P, Maas P, Wilcox A, Wheeler W, Brook M, Check D, et al. iCARE: An R package to build, validate and apply absolute risk models. PloS one. 2020;15(2):e0228198.

47. Database HSaHR. 2020.

48. Organization WH. Global Health Observatory indicator views 2018 [Available from: https://apps.who.int/gho/data/node.imr.LIFE_0000000029?lang=en.

49. Ferlay J, Colombet M, Soerjomataram I, Mathers C, Parkin DM, Pineros M, et al. Estimating the global cancer incidence and mortality in 2018: GLOBOCAN sources and methods. International journal of cancer. 2019;144(8):1941–53.

50. Naber SK, Kundu S, Kuntz KM, Dotson WD, Williams MS, Zauber AG, et al. Cost-Effectiveness of risk-stratified colorectal cancer screening based on polygenic risk: current status and future potential. JNCI cancer spectrum. 2020;4(1):pkz086.

51. Schmit SL, Edlund CK, Schumacher FR, Gong J, Harrison TA, Huyghe JR, et al. Novel Common Genetic Susceptibility Loci for Colorectal Cancer. Journal of the National Cancer Institute. 2019;111(2):146–57.

52. Armaroli P, Villain P, Suonio E, Almonte M, Anttila A, Atkin WS, et al. European Code against Cancer, 4th Edition: Cancer screening. Cancer epidemiology. 2015;39 Suppl 1:S139–52.

53. Autier P. Personalised and risk based cancer screening. BMJ (Clinical research ed). 2019;367.

54. Doubeni C. Screening for colorectal cancer: Strategies in patients at average risk 2019 [Available from: https://www.uptodate.com/contents/screening-for-colorectal-cancer-strategies-inpatients-at-average-risk.

55. Iwasaki M, Tanaka-Mizuno S, Kuchiba A, Yamaji T, Sawada N, Goto A, et al. Inclusion of a genetic risk score into a validated risk prediction model for colorectal cancer in Japanese men improves performance. Cancer prevention research. 2017;10(9):535–41.

56. Weigl K, Thomsen H, Balavarca Y, Hellwege JN, Shrubsole MJ, Brenner H. Genetic Risk Score Is Associated With Prevalence of Advanced Neoplasms in a Colorectal Cancer Screening Population. Gastroenterology. 2018;155(1):88–98.e10.

57. Weigl K, Chang-Claude J, Knebel P, Hsu L, Hoffmeister M, Brenner H. Strongly enhanced colorectal cancer risk stratification by combining family history and genetic risk score. Clinical epidemiology. 2018;10:143.

58. Archambault AN, Su Y-R, Jeon J, Thomas M, Lin Y, Conti DV, et al. Cumulative Burden of Colorectal Cancer–Associated Genetic Variants Is More Strongly Associated With Early-Onset vs Late-Onset Cancer. Gastroenterology. 2020;158(5):1274–86. e12.

59. Oncology ESfM. Decline in colorectal cancer deaths in Europe is a ‘major success’ story 2018 [Available from: https://www.eurekalert.org/pub_releases/2018-03/esfm-dic031518.php.

60. Hall N, Birt L, Rees CJ, Walter FM, Elliot S, Ritchie M, et al. Concerns, perceived need and competing priorities: a qualitative exploration of decision-making and non-participation in a population-based flexible sigmoidoscopy screening programme to prevent colorectal cancer. BMJ open. 2016;6(11):e012304.

61. Issa IA, Noureddine M. Colorectal cancer screening: An updated review of the available options. World journal of gastroenterology. 2017;23(28):5086.

62. Li B, Gan A, Chen X, Wang X, He W, Zhang X, et al. Diagnostic Performance of DNA Hypermethylation Markers in Peripheral Blood for the Detection of Colorectal Cancer: A Meta-Analysis and Systematic Review. PloS one. 2016;11(5):e0155095.

63. Lieberman DA, Rex DK, Winawer SJ, Giardiello FM, Johnson DA, Levin TR. Guidelines for colonoscopy surveillance after screening and polypectomy: a consensus update by the US Multi-Society Task Force on Colorectal Cancer. Gastroenterology. 2012;143(3):844–57.

